# October 2018 – December 2023 time-series analysis of pediatric RSV immunizations and RSV-associated hospitalizations

**DOI:** 10.1101/2024.01.25.24301780

**Authors:** Brianna M. Goodwin Cartwright, Samuel Gratzl, Patricia J Rodriguez, Charlotte Baker, Nick Stucky

## Abstract

This study describes two population under age two 1) who received an RSV immunization and 2) experienced RSV-associated hospitalizations since 2018. Results show low uptake of the RSV immunization. RSV-associated hospitalizations exhibited earlier and higher peaks in the 2021/22 and 2022/23 seasons compared to previous years.

---

Respiratory syncytial virus (RSV) is an infectious respiratory virus, which typically spreads between October and February (Hamid et al., 2023). The seasonality of RSV was disrupted during the COVID-19 pandemic, likely due to social behavioral factors associated with COVID; this lead to an absent 2021 season and an earlier and longer season in 2022 (Hamid et al., 2023). The summer 2022 RSV peak particularly affected a pediatric population (Bourdeau et al., 2023). However, it is unknown how this translates to hospitalizations for this population.

Between May and August of 2023, the FDA approved RSV immunizations for people over age 60 (RSVPreF3 or arexvy) (U.S. Food & Drug Administration, 2023a), children under 24 months of age (nirsevimab or beyfortus) (U.S. Food & Drug Administration, 2023c), and pregnant women (RSVpreF or abrysvo) to prevent illness in infants (U.S. Food & Drug Administration, 2023b). These new treatment options added to the existing monoclonal antibody treatment available for infants. However, little data exists on the uptake of these immunizations, specifically in the pediatric population. Further, given the rise in RSV-associated infections in pediatric populations, little data are available to understand trends in recent RSV-associated hospitalizations.

In this study we aimed to 1) describe the rate and demographics of children under age two who received an RSV immunization between October 2018 and December 2023 and 2) describe the weekly-rate of pediatric RSV-associated hospitalizations.

## Methods

This study used a subset of Truveta Data. Truveta provides access to continuously updated and de-identified electronic health record (EHR) data from a collective of US health care systems, including structured information on demographics, encounters, diagnoses, vital signs, medications, and laboratory and diagnostic tests. Data are normalized into a common data model through syntactic and semantic normalization. Truveta Data are then de-identified by expert determination under the HIPAA Privacy Rule. Data for this study were accessed on January 19, 2024.

Using these data, we conducted two analyses. First, we identified the rate of eligible children receiving an RSV immunization. Second, we compared yearly seasonal RSV-associated hospitalization trends.

### *Rate of eligible children receiving an RSV* immunization

We included children who had received a previous DTaP, Influenza, Hepatitis B, Pneumococcal pneumonia, or Rotavirus immunization between October 2018 and December 2023. Children were considered eligible for the RSV immunization on a rolling-basis, for the 12 months after they received one of the prior immunizations or until they received an RSV immunization (Hamid et al., 2023). If a child received a subsequent immunization, the 12-month period restarted.

Each month children were grouped into six-month age groups (0-6 months, 6-12 months, 12-18 months, and 18-24 months). We identified the month that children received their first RSV immunization and the age group designation at the time of their immunization. If a child received more than one RSV immunization, only the first was counted. We then calculated the monthly rate of first-time RSV immunization administrations per eligible population.

All immunizations were defined using CVX and CPT codes (available in the supplement).

### Yearly RSV-associated hospitalization trends

We included children between one week and under two years of age, who had been hospitalized between October 2018 and December 2023. We used a minimum age of one week to exclude hospital stays coinciding with a child’s birth. Within this population we identified children who tested positive for RSV within 14 days before or after the start of their hospitalization. RSV seasons were defined as October to September of the subsequent year.

We calculated the weekly rate of RSV-associated hospitalizations compared to all hospitalizations.

## Results

### Rate of eligible children receiving an RSV immunization

We included 1,872,292 children under age two who were eligible to receive an RSV immunization. Within this population, 0.9% received an RSV immunization between October 2018 and December 2023. Since the immunization was approved in July 2023, 3.7% of the eligible population received an immunization, with higher rates occurring in the youngest age groups (0-6 months: 4.9%, 6-12 months: 1.9%, 12-18 months: 0.6%, and 18-24 months: 0.2%). The population who received an RSV immunization was primarily male (51.7%), white (48.3%), and not Hispanic or Latino (64.7%). Additional demographic descriptions are available in the supplement. The population receiving an RSV immunization was also primarily under six months of age (80.4%), while they only made up 46.1% of the eligible population. Most RSV immunizations occurred between October and December of 2023 (Figure 1A).

**Figure 1:**
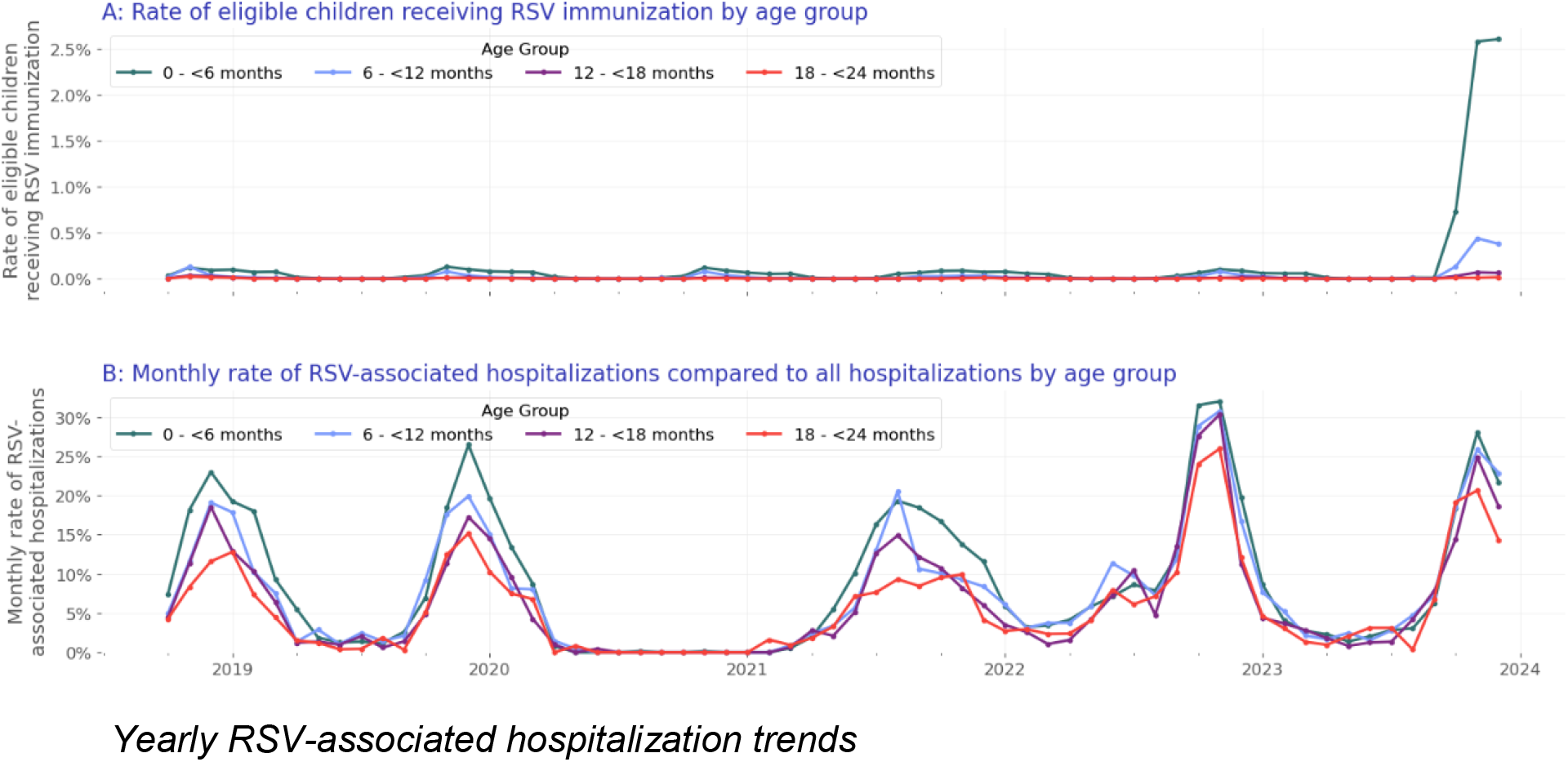
A) The rate of first-time RSV immunization per eligible population per age group and B) The weekly rate of RSV-associated hospitalizations compared to all hospitalizations for children under 2 years of age.

We included 93,228 children under age two who were hospitalized (for any reason) between October 2018 and December 2023. Of those, 13.9% (12,939) were RSV-associated hospitalizations. The population with RSV-associated hospitalizations were primarily between 0-6 months of age (54.6%), male (56.4%), white (50.0%) and not Hispanic or Latino (63.8%). Additional demographic descriptions are available in the supplement.

Prior to the COVID pandemic, in the 2018/2019 and 2019/2020 seasons, the rate of RSV-associated hospitalizations began to increase in November and subsided by March/April of the following year (Figure 1B). In the 2020/2021 season, RSV-associated hospitalizations were rare during the typical periods of high hospitalizations (October 2021 – March 2022); however, the 2021/2022 seasonal increase started earlier than normal, namely at the end of the previous respiratory virus season (July 2021). The 2022/2023 season saw both earlier (rates above 10% starting in September) and higher RSV-associated hospitalization rates compared to prior years (2022 seasonal peak of 30.7% of hospitalizations, compared to 2020 seasonal peak of 17.2% hospitalizations). During the 2023/2024 season, rates began to increase in October (with a rate of 17.8%), and the current seasonal peak occurred at 26.2% in November 2023.

Although the 0–6-month-old group comprises the largest percent of hospitalizations across all seasons, more recent seasons see an increasing rate of RSV-associated hospitalizations in all older age groups; the 6–12-month age group increased from 20.0% of the RSV-associated hospitalized population in the 2018/2019 season to 22.8% of the population in the current 2023/2024 season. We also see this increasing trend in the 12–18-month-old age group and 18 – 24-month-old age group, with rates increasing from 14.1% to 15.6% and 7.9% to 9.9%, respectively.

## Discussion

Surveillance trends on positive detection of RSV exist and show similar seasonal patterns to our study (Hamid et al., 2023). However, little data exists on both the uptake in pediatric RSV immunizations now that they are available and seasonal trends of pediatric hospitalizations.

This report is one of the first to demonstrate low uptake in pediatric RSV immunization (less than 4% of the eligible population since approval in July 2023). RSV immunizations may be low in this population due to higher demand than supply of the immunization (Sanofi, 2023). This report also highlights the importance of increased immunizations with the return to seasonal pediatric RSV trends and higher November 2022 and 2023 rates, than prior peak seasonal rates (through 2018). We also see an increasing percentage of RSV-associated hospitalizations for children over 6 months old. This is consistent with Rao and colleagues (Rao et al., 2023) and highlights the continued need for immunizations in the older age groups, as well as those in the 0-6 month-old age group.

There are limitations associated with this study. First, an RSV vaccine became available in August 2023 for women during the 32-36 weeks of pregnancy to protect their babies after birth. We did not exclude children whose mothers received this vaccine. However, this population is likely small given the timeline of vaccine rollout. Further, this would only affect the youngest age group (0-6-month-olds), which is the largest immunized population here. Second, children were only considered eligible for the immunization if they had previously received an immunization.

Since the data used in this study are EHR data, this assumption was made to ensure we did not miss children who were immunized outside a Truveta health system. This may inflate the percentage of the population who received an immunization. Finally, children were considered to have an RSV-associated hospitalization if they tested positive for RSV, independent of the reason they were admitted.

Despite these limitations, this study adds knowledge about the percentages and distributions of pediatric populations receiving RSV immunizations and the RSV-associated hospitalization trends between October 2018 and December 2023.

## Data Availability

The data used in this study are available to all Truveta subscribers and may be accessed at studio.truveta.com.

