## Supplemental Tables for "October 2018 – December 2023 time-series analysis of pediatric RSV immunizations and RSV-associated hospitalizations"

Supplement

Table 1: Code sets used to define each immunization

| Data Def | System | Code |
| --- | --- | --- |
| DTaP | CPT | 90700, 90696, 90721, 90723 |
|  | CVX | 107, 120, 50, 20, 110, 146, 170, 106, 132, 130 |
| Influenza | CPT | 90674, 90672, 90724, 90667, 90685, 90662, 90686, 90655, 90687, 90756, 90658, 90630, 90694, 90653, 90673, 90688, 90654, 90682, 90656, 90689, 90657 |
|  | CVX | 168, 135, 155, 149, 158, 144, 205, 197, 166, 141, 185, 171, 88, 161, 186, 150, 140 |
| Hepatitis B | CPT | 90744, 90748, 90746, 90739, 90740, 90747, 90743, 90723, 90636, 90745, 90697, 90759 |
|  | CVX | 220, 102, 42, 44, 104, 189, 110, 146, 08, 132, 198, 45, 193, 43, 51 |
| Rotavirus | CPT | 90681, 90680 |
|  | CVX | 74, 116, 122, 119 |
| Pneumococcal | CPT | 90732, 90677, 90671, 90669, 90670 |
|  | CVX | 215, 133, 216, 177, 100, 109, 33, 152 |
| RSV | CPT | 90679, 90678 |
|  | CVX | 145, 307, 93, 314, 305, 306, 304, 315, 71, 303 |

Table 2: Distribution of population who was eligible to receive an RSV immunization and those who received an RSV immunization during the study period.

|  |  | Eligible population | RSV immunized population |
| --- | --- | --- | --- |
|  |  | 1,872,292 (100.0%) | 17,600 (100.0%) |
| Age | 0 - <6 months | 862,670 (46.1%) | 14,156 (80.4%) |
|  | 12 - <18 months | 195,842 (10.5%) | 439 (2.5%) |
|  | 18 - <24 months | 229,983 (12.3%) | 154 (0.9%) |
|  | 6 - <12 months | 583,797 (31.2%) | 2,851 (16.2%) |
| Sex | Female | 908,416 (48.5%) | 8,441 (48.0%) |
|  | Male | 959,403 (51.2%) | 9,100 (51.7%) |
|  | Unknown | 4,473 (0.2%) | 59 (0.3%) |
| Race | American Indian or Alaska Native | 9,869 (0.5%) | 128 (0.7%) |
|  | Asian | 122,159 (6.5%) | 1,113 (6.3%) |
|  | Black or African American | 217,831 (11.6%) | 2,542 (14.4%) |
|  | Native Hawaiian or Other Pacific Islander | 20,724 (1.1%) | 334 (1.9%) |
|  | Other Race | 200,123 (10.7%) | 1,522 (8.6%) |
|  | Unknown | 361,631 (19.3%) | 3,460 (19.7%) |
|  | White | 939,955 (50.2%) | 8,501 (48.3%) |
| Ethnicity | Hispanic or Latino | 404,238 (21.6%) | 2,799 (15.9%) |
|  | Not Hispanic or Latino | 1,096,771 (58.6%) | 11,386 (64.7%) |
|  | Unknown | 371,283 (19.8%) | 3,415 (19.4%) |

Table 3: Distribution of population who was hospitalized overall, and specifically for those with an RSV-associated hospitalization since October 2018.

|  |  | Hospitalized population | RSV-associated hospitalization population |
| --- | --- | --- | --- |
|  |  | 93,228 (100.0%) | 12,939 (100.0%) |
| Age | 0 - <6 months | 45,011 (48.3%) | 7,065 (54.6%) |
|  | 12 - <18 months | 16,300 (17.5%) | 1,923 (14.9%) |
|  | 18 - <24 months | 13,575 (14.6%) | 1,257 (9.7%) |
|  | 6 - <12 months | 18,342 (19.7%) | 2,694 (20.8%) |
| Sex | Female | 39,845 (42.7%) | 5,618 (43.4%) |
|  | Male | 52,881 (56.7%) | 7,300 (56.4%) |
|  | Unknown | 502 (0.5%) | 21 (0.2%) |
| Race | American Indian or Alaska Native | 952 (1.0%) | 115 (0.9%) |
|  | Asian | 5,856 (6.3%) | 705 (5.4%) |
|  | Black or African American | 14,235 (15.3%) | 1,903 (14.7%) |
|  | Native Hawaiian or Other Pacific Islander | 2,841 (3.0%) | 351 (2.7%) |
|  | Other Race | 15,618 (16.8%) | 2,334 (18.0%) |
|  | Unknown | 9,305 (10.0%) | 1,064 (8.2%) |
|  | White | 44,421 (47.6%) | 6,467 (50.0%) |
| Ethnicity | Hispanic or Latino | 24,628 (26.4%) | 3,540 (27.4%) |
|  | Not Hispanic or Latino | 58,207 (62.4%) | 8,254 (63.8%) |
|  | Unknown | 10,393 (11.1%) | 1,145 (8.8%) |
